# Relationships of serum osmolality with cardiovascular disease and mortality risk: A retrospective cohort study of the National Health and Nutrition Examination Surveys database

**DOI:** 10.1101/2025.09.22.25336381

**Authors:** Kun Cao, Feng Hai

## Abstract

**Aim:** The relationship of serum osmolality with cardiovascular disease (CVD) is unclear. Serum osmolality calculated by sodium (Na), potassium (K), glucose, urea nitrogen and other indicators has been proposed to replace the complex and high cost directly measured plasma osmolality (pOsm). Herein, we aimed to investigate the association of calculated serum osmolality with CVD in adults, and further assess the mortality risk in patients with CVD.

**Methods:** In this retrospective cohort study, data of 28,715 adults were extracted from the National Health and Nutrition Examination Surveys (NHANES) database in 2007-2018. We explored the association of serum osmolality with CVD in adults using weighted univariate and multivariate logistic regression analyses, and further investigated relationship of serum osmolality with all-cause mortality in patients with CVD through weighted Cox regression analyses. The evaluation indexes were odds ratios (ORs), hazard ratios (HRs) and 95% confidence intervals (CIs). These relationships were also explored in subgroups of age, gender, stroke, cancer, hypertension, and diabetes mellitus (DM).

**Results:** A total of 2,415 (6.75%) individuals had CVD, and 706 (24.40%) of them died from all-cause. After adjusting for covariates respectively, we found that current dehydration (serum osmolality >300 mmol/L) was associated with both high odds of CVD (OR=1.26, 95%CI: 1.02-1.57) and high risk of all-cause mortality (OR=1.41, 95%CI: 1.13-1.75). In addition, the relationships between current dehydration and CVD were also significant in ≥60 years old, male, non-stroke, non-cancer, hypertension, and non-DM subgroups (all *P*<0.05).

**Conclusion:** Serum osmolality is important for adults especially when they at dehydration status, which may be associated with potential risk of CVD. Besides, patients with CVD should also pay more attention to the current dehydration to reduce the risk of mortality.

## Introduction

Cardiovascular disease (CVD) is a leading cause of death and disability worldwide [1]. Despite a decline in age-standardized CVD mortality globally over the past 30 years, epidemiological surveys have shown that CVD still accounted for one-third of all mortality worldwide in 2019 [2]. Besides, CVD has been recognized as a preventable disease because the modifiable risk factors account for over 90% of the risk for developing CVD [3]. Therefore, exploring preventative measures that reduce CVD risk is of great necessary to lessen the burden caused by CVDs.

Serum osmolality is the sum concentrations of every single dissolved particle in the blood, and is strongly influenced by sodium (Na), potassium (K), glucose, and urea [4]. Serum osmolality is essential to maintain the physiological function of various organs, and dehydration reflected by hyperosmosis is considered to be an important factor related to human health [5, 6]. Hyperosmosis increases the level of proinflammatory factors in vivo [7], and serum osmolality can cause changes in cell morphology that may destroy key components within the cell and eventually lead to cell death [8]. The directly measured plasma osmolality (pOsm) is recognized as the golden standard to reflect hydration status, and however, due to the complexity and high cost of the operation, pOsm determination is usually not a routine inspection item. Based on this, serum osmolality calculated by Na, k, glucose, urea nitrogen and other indicators has been proposed to replace direct measurement, among which the European Society of Enteral and Parenteral (ESPEN) equation has proved to be highly accurate, and is recommended as a marker for assessment of dehydration in clinical practice [5, 9]. Previous studies suggested hydration status was significantly associated with the risk of metabolic syndrome (MS) and stroke [10, 11]. In addition, osmolality is also linked to the risk of mortality in both the general population and patients with stroke [12, 13]. Nevertheless, no study has investigated the relationship of serum osmolality with CVD and the risk of mortality in patients with CVD.

Herein, the current research aims to explore the association of serum osmolality with CVD risk in general population basing on the National Health and Nutrition Examination Surveys (NHANES) database, and further assessed the role of serum osmolality in mortality among patients with CVD. We hope the study findings may provide some references for exploration on clinical index related to CVD prevention.

## Methods

### Study design and participants

This is a retrospective cohort study. Data of adults (aged 20 years old) were extracted from the NHANES database in 2007-2018. The NHANES survey is jointly conducted by the Centers for Disease Control and Prevention (CDC) and the National Center for Health Statistics (NCHS), which aims to assess nutritional and health status of noninstitutionalized population in the United States. More details are shown elsewhere: https://www.cdc.gov/nchs/nhanes/index.htm. The NHANES includes a complex, multistage stratified probability sample based on selected counties, blocks, households, and persons within households. Interviews in participants’ homes were conducted by the NCHS well trained professionals, and extensive physical examinations were conducted at mobile exam centers (MECs).

Initially, 34,760 adults with information on CVD assessment were included. The exclusion criteria were: (1) missing information on examination of glucose, Na, K, or blood urea nitrogen (BUN), (2) missing survival data, and (3) missing information on study variables including hypertension, dyslipidemia, cancer, stroke, estimated glomerular filtration rate (eGFR), physical activity, smoking, body mass index (BMI), or total energy intake. A total of 28,715 were eligible. The institutional review board (IRB) of the NCHS has approved the NHANES. Since study data were publicly available and de-identified, and informed consent has been obtained from the participants, no ethical approval of our agency’s IRB was required.

### Measurement of serum osmolality

In the NHANES, participants’ blood samples were collected through physical examinations by trained professionals at MECs. Then, the blood samples were divided into aliquots and shipped to multiple laboratories for analysis. The serum osmolality was calculated via the formula [14]: osmolarity = 1.86 × (Na + K) + 1.15 × glucose + BUN + 14. According to the serum osmolarity level, individuals were divided into four statuses [9, 15]: hyperhydration (<285 mmol/L), normal hydration (285-294 mmol/L), impending dehydration (295-300 mmol/L), and current dehydration (>300 mmol/L).

### Study outcomes and follow-up time

The study outcomes were CVD and all-cause mortality. Information on CVD was collected through the NHANES multiple-choice question (MCQ) with the question: “Have you ever been told you had (congestive) heart failure, coronary heart disease, angina/angina pectoris, heart attack, stroke.” Person self-reported having any one disease was recognized as a patient with CVD [16]. The mortality status of the patients was determined through the NHANES publicly linked mortality file, which was correlated with the NCHS with the National Death Index (NDI) through a probability matching algorithm. For more details, please visit their website: https://ftp.cdc.gov/pub/health_statistics/NCHS/datalinkage/linked_mortality/. The follow-up ended at December 31, 2019.

### Variables extraction

We also included the following study variables: age, gender, race, poverty income ratio (PIR), marital status, education level, height, weight, BMI, physical activity, smoking, stroke, cancer, hypertension, diabetes mellitus (DM), dyslipidemia, cardiovascular agents, total energy intake, Alternate Mediterranean Diet (aMed) score, and eGFR.

Basing on the world health organization (WHO) criteria, BMI were divided into four levels, including <25 kg/m^2^, [25, 30) kg/m^2^, and ≥30 kg/m^2^. In formation on physical activity was collected via the physical activity questionnaire (PAQ) in NHANES, and was converted to metabolic equivalent (MET). The calculation formula was: Energy expenditure (MET·min) = recommended MET × exercise time of corresponding activity (min). Physical activity was divided into two levels with the cut-off value of 450 MET·min/week [17]. Persons self-reported to have smoked at least 100 cigarettes in their lives were considered as smokers, which contained former smoker (who have quitted) and current smoker. Hypertension was diagnosed on the basis of self-reported high blood pressure, currently took medication for high blood pressure, or a measured systolic blood pressure (SBP) ≥130 mm Hg or diastolic blood pressure (DBP) ≥80 mm Hg. DM was defined according to self-reported usage of oral hypoglycemic agents or insulin, glycosylated hemoglobin (HbAlc) ≥6.5%, a plasma glucose level ≥200 mg/dL at 2 hours after the oral glucose tolerance test, or a fasting glucose level ≥126 mg/dL. Patients with total cholesterol (TC) ≥200 mg/dL (5.2 mmol/L) or triglycerides (TG) ≥150 mg/dL (1.7 mmol/L) or low-density lipoprotein cholesterol (LDL-C) ≥130 mg/dL (3.4 mmol/L) or high-density lipoprotein cholesterol (HDL-C) ≤40 mg/dL (1.0 mmol/L) or self-reported hypercholesteremia or receiving lipid-lowering therapy were identified as dyslipidemia [18]. Cancer was diagnosed through positive answer to the question: “Have you ever been told you had (congestive) cancer or malignancy.” eGFR was calculated through the formula: eGFR_CKD-EPI_ (mL/min/1.73m^2^) = 141 × min (*Scr*/*κ*,1) ^α^ × max (*Scr*/*κ*,1)^-1.029^ × 0.993^age^ × 1.108 (if female) × 1.159 (if Black), where κ is 0.7 for females and 0.9 for males, α is −0.329 for females and −0.411 for males, min indicates the minimum of Scr/κ or 1, and max indicates the maximum of Scr/κ or 1, and the unit of Scr is mg/dL [19]. The eGFR was divided into two levels with the cut-off value 60. In addition, the aMed score (total score=18) was derived by an assigned value of “0”, “1”, or “2” across nine food categories (including vegetables, legumes, fruits, nuts, whole grains, red and processed meats, fish, alcohol, and olive oil), with higher scores indicating better adherence to the MedDiet pattern [20, 21].

### Statistical analysis

Normally distributed data were described with mean ± standard error (mean ± SE), weighted analysis of variance (ANOVA) was used for comparation among different group. Categorical data were expressed by frequency with constituent ratio [N (%)], and chi-square test (χ²) was used for comparisons. We used the NHANES special sample weights: MEC two-year sample weights (WTMEC2YR) for analyses.

When CVD as the study outcome, weighted univariate logistic regression analysis was employed to screen covariate, and weighted univariate and multivariate logistic regression analyses were used to explored the association of serum osmolality with CVD in general populations. Model 1 adjusted for demographic variables including age, gender, race, PIR, marital status, and education level. Model 2 additionally adjusted for BMI, physical activity, smoking, stroke, cancer, hypertension, DM, dyslipidemia, cardiovascular agents, total energy intake, and eGFR.

When all-cause mortality as the study outcome, we used weighted univariate Cox regression analysis for covariates screening, and used weighted univariate and multivariate Cox regression analyses to investigate the relationship of serum osmolality with all-cause mortality in patients with CVD. Model 1 adjusted for demographic variables including age, race, marital status, and education level. Model 2 adjusted for physical activity, smoking, stroke, cancer, hypertension, DM, cardiovascular agents, aMed score, and eGFR in addition to Model 1.

We further investigated these two relationships in subgroups of age, gender, stroke, cancer, hypertension, and DM. In addition, Kaplan-Meier (KM) curve on the survival probability in persons with different hydration statuses. The evaluation indexes were odds ratios (ORs) and hazard ratios (HRs) with 95% confidence intervals (CIs) respectively. Two-sided *P*<0.05 was considered significant. Statistical analysis was performed by R version 4.2.3 (Institute for Statistics and Mathematics, Vienna, Austria).

## Results

### Characteristics of participants

The process of participants screening is shown in the Figure 1. There were 34,760 adults with information on CVD in the NHANES database in 2007-2018. Those who missing information on glucose, Na, K, or BUN (n=3498), survival (n=71), hypertension (n=1), dyslipidemia (n=1), cancer (n=24), stroke (n=33), eGFR (n=1), physical activity (n=4), smoking (n=22), BMI (n=387), or total energy intake (n=2003). Finally, 28,715 were eligible.

**Figure 1.**
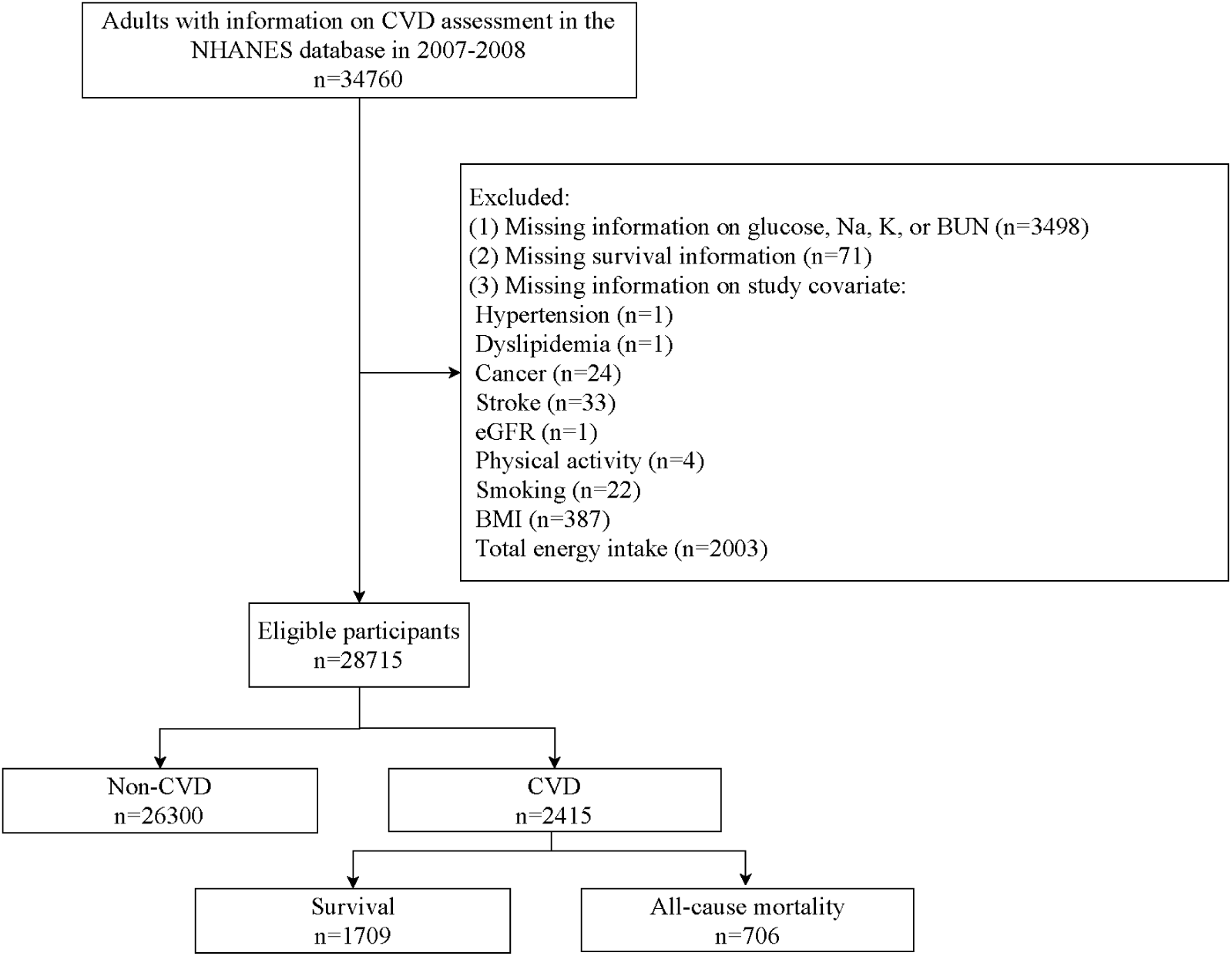
Flowchart of participants screening.

The characteristics of adults among different hydration status groups are shown in the Table 1. The average age of total participants was 47.48 years old, and that in current dehydration group (60.47 years old) was the highest, followed by that in impending dehydration group (54.26 years old). Serum osmolality as well as its related indexes were all different among different hydration status groups (all *P*<0.001). A total of 2,415 (6.75%) persons had CVD, and within them, 706 (24.40%) died from all-cause. In addition, gender, race, PIR, marital status, education level, height, weight, BMI, physical activity, smoking, stroke, cancer, hypertension, DM, dyslipidemia, cardiovascular agents, total energy intake, and eGFR were all significantly among these four groups (all *P*<0.05).

**Table 1.** Characteristics of eligible adults among different hydration statuses groups.

| Variables | Total<br>(n=28715) | Normal<br>hydration<br>(n=18696) | Impending<br>dehydration<br>(n=5680) | Current<br>dehydration<br>(n=2117) | Hyperhydration<br>(n=2222) | Statistics | *P* |
| --- | --- | --- | --- | --- | --- | --- | --- |
| Age, years, Mean ± SE | 47.48 ± 0.24 | 44.96 ± 0.26 | 54.26 ± 0.62 | 60.47 ± 0.67 | 42.99 ± 0.53 | F=220.747 | <0.001 |
| Age, years, n (%) | | | | | | $\chi^2=310.323$ | <0.001 |
| <60 | 19127 (73.68) | 13929 (79.86) | 2757 (58.00) | 691 (41.79) | 1750 (82.63) |  |  |
| $\geq 60$ | 9588 (26.32) | 4767 (20.14) | 2923 (42.00) | 1426 (58.21) | 472 (17.37) | | |
| Gender, n (%) | | | | | | $\chi^2=73.437$ | <0.001 |
| Male | 13982 (48.40) | 9041 (47.97) | 3094 (55.89) | 1179 (54.36) | 668 (30.13) |  |  |
| Female | 14733 (51.60) | 9655 (52.03) | 2586 (44.11) | 938 (45.64) | 1554 (69.87) |  |  |
| Race, n (%) | | | | | | $\chi^2=2.680$ | 0.016 |
| Non-Hispanic White | 12129 (67.53) | 7645 (66.52) | 2543 (70.12) | 956 (69.99) | 985 (68.01) |  |  |
| Non-Hispanic Black | 5902 (10.56) | 3884 (10.78) | 1074 (9.07) | 448 (10.47) | 496 (12.33) |  |  |
| Mexican American | 4408 (8.55) | 2900 (8.75) | 898 (8.42) | 312 (7.90) | 298 (7.63) |  |  |
| Other races | 6276 (13.36) | 4267 (13.95) | 1165 (12.38) | 401 (11.64) | 443 (12.04) |  |  |
| PIR, n (%) | | | | | | $\chi^2=6.730$ | <0.001 |
| <1 | 5566 (13.19) | 3706 (13.52) | 969 (11.03) | 381 (11.72) | 510 (16.56) |  |  |
| $\geq 1$ | 20631 (79.76) | 13444 (79.79) | 4143 (80.66) | 1506 (79.72) | 1538 (77.43) | | |
| Unknown | 2518 (7.06) | 1546 (6.70) | 568 (8.31) | 230 (8.56) | 174 (6.01) |  |  |
| Marital status, n (%) | | | | | | $\chi^2=4.561$ | 0.006 |
| Married/living with partner | 17147 (63.80) | 11216 (64.02) | 3500 (65.54) | 1206 (61.50) | 1225 (59.65) |  |  |
| Single/separated/divorced/widowed | 11568 (36.20) | 7480 (35.98) | 2180 (34.46) | 911 (38.50) | 997 (40.35) |  |  |
| Education level, n (%) | | | | | | $\chi^2=3.954$ | 0.003 |
| Below high school | 6895 (15.41) | 4299 (14.85) | 1484 (15.74) | 615 (18.15) | 497 (17.23) |  |  |
| High school/GED or equivale | 6572 (23.06) | 4235 (22.64) | 1327 (24.19) | 520 (26.67) | 490 (21.29) |  |  |
| Above high school | 15248 (61.53) | 10162 (62.51) | 2869 (60.07) | 982 (55.18) | 1235 (61.48) |  |  |
| Height, cm, Mean ± SE | 168.60 ± 0.10 | 168.82 ± 0.12 | 169.10 ± 0.22 | 167.77 ± 0.26 | 166.21 ± 0.27 | F=31.971 | <0.001 |
| Weight, kg, Mean ± SE | 83.03 ± 0.24 | 82.62 ± 0.28 | 85.35 ± 0.41 | 86.22 ± 0.65 | 78.66 ± 0.67 | F=31.411 | <0.001 |
| BMI, kg/m<sup>2</sup>, n (%) | | | | | | $\chi^2=16.713$ | <0.001 |
| <25 | 8076 (29.14) | 5493 (30.49) | 1364 (24.27) | 456 (22.01) | 763 (34.63) |  |  |
| [25, 30) | 9484 (33.01) | 6229 (33.18) | 1908 (34.14) | 674 (30.03) | 673 (31.14) |  |  |
| $\geq 30$ | 11155 (37.85) | 6974 (36.33) | 2408 (41.59) | 987 (47.96) | 786 (34.23) | | |
| Physical activity, MET·min/week, n (%) | | | | | | $\chi^2=34.968$ | <0.001 |
| <450 | 7591 (21.99) | 4479 (20.22) | 1689 (23.89) | 826 (34.40) | 597 (22.99) |  |  |
| ≥450 | 21124 (78.01) | 14217 (79.78) | 3991 (76.11) | 1291 (65.60) | 1625 (77.01) |  |  |
| Smoking, n (%) | | | | | | $\chi^2=24.573$ | <0.001 |
| Never | 15991 (55.74) | 10574 (56.57) | 3054 (54.36) | 1123 (54.46) | 1240 (53.03) |  |  |
| Former | 6936 (24.91) | 4084 (23.11) | 1717 (30.33) | 728 (33.43) | 407 (20.92) |  |  |
| Now | 5788 (19.34) | 4038 (20.32) | 909 (15.31) | 266 (12.11) | 575 (26.05) |  |  |
| Stroke, n (%) | | | | | | $\chi^2=59.399$ | <0.001 |
| No | 27641 (97.24) | 18197 (97.96) | 5377 (96.25) | 1907 (91.52) | 2160 (97.80) |  |  |
| Yes | 1074 (2.76) | 499 (2.04) | 303 (3.75) | 210 (8.48) | 62 (2.20) |  |  |
| Cancer, n (%) | | | | | | $\chi^2=54.597$ | <0.001 |
| No | 25992 (89.82) | 17301 (91.70) | 4893 (85.00) | 1740 (81.48) | 2058 (91.74) |  |  |
| Yes | 2723 (10.18) | 1395 (8.30) | 787 (15.00) | 377 (18.52) | 164 (8.26) |  |  |
| Hypertension, n (%) | | | | | | $\chi^2=125.054$ | <0.001 |
| No | 12642 (48.71) | 9323 (53.74) | 1779 (37.11) | 391 (23.75) | 1149 (52.54) |  |  |
| Yes | 16073 (51.29) | 9373 (46.26) | 3901 (62.89) | 1726 (76.25) | 1073 (47.46) |  |  |
| DM, n (%) | | | | | | $\chi^2=352.827$ | <0.001 |
| No | 23494 (86.40) | 16379 (90.60) | 4070 (78.21) | 1026 (56.90) | 2019 (92.59) |  |  |
| Yes | 5221 (13.60) | 2317 (9.40) | 1610 (21.79) | 1091 (43.10) | 203 (7.41) |  |  |
| Dyslipidemia, n (%) | | | | | | $\chi^2=49.166$ | <0.001 |
| No | 8332 (30.21) | 6016 (32.91) | 1194 (22.94) | 317 (17.32) | 805 (34.32) |  |  |
| Yes | 20383 (69.79) | 12680 (67.09) | 4486 (77.06) | 1800 (82.68) | 1417 (65.68) |  |  |
| Cardiovascular agents, n (%) | | | | | | $\chi^2=163.611$ | <0.001 |
| No | 25180 (89.69) | 17034 (92.27) | 4629 (84.15) | 1484 (74.42) | 2033 (92.55) |  |  |
| Yes | 3535 (10.31) | 1662 (7.73) | 1051 (15.85) | 633 (25.58) | 189 (7.45) |  |  |
| Total energy intake, kcal, Mean ± SE | 2180.34 ± 8.22 | 2208.98 ± 9.74 | 2166.61 ± 18.76 | 2053.38 ± 32.94 | 2069.41 ± 25.99 | F=13.969 | <0.001 |
| aMed score, Mean ± SE | 4.72 ± 0.03 | 4.69 ± 0.03 | 4.81 ± 0.06 | 4.86 ± 0.08 | 4.69 ± 0.08 | F=2.441 | 0.069 |
| eGFR, mL/min/1.73m<sup>2</sup>, n (%) | | | | | | $\chi^2=424.010$ | <0.001 |
| <60 | 1640 (4.07) | 421 (1.75) | 548 (7.30) | 642 (24.05) | 29 (0.94) |  |  |
| $\geq 60$ | 27075 (95.93) | 18275 (98.25) | 5132 (92.70) | 1475 (75.95) | 2193 (99.06) | | |
| Serum osmolarity, mmol/L, Mean ± SE | 291.84 ± 0.17 | 290.49 ± 0.05 | 297.01 ± 0.04 | 303.56 ± 0.12 | 282.25 ± 0.09 | F=9706.098 | <0.001 |
| Glucose, mmol/L, Mean ± SE | 5.53 ± 0.02 | 5.27 ± 0.02 | 6.03 ± 0.05 | 7.63 ± 0.17 | 4.94 ± 0.03 | F=139.153 | <0.001 |
| Na, mmol/L, Mean ± SE | 139.33 ± 0.09 | 138.98 ± 0.04 | 141.22 ± 0.07 | 142.64 ± 0.18 | 135.35 ± 0.05 | F=1696.964 | <0.001 |
| K, mmol/L, Mean ± SE | 4.00 ± 0.01 | 3.96 ± 0.00 | 4.11 ± 0.01 | 4.29 ± 0.02 | 3.83 ± 0.01 | F=247.741 | <0.001 |
| BUN, mmol/L, Mean ± SE | 4.89 ± 0.02 | 4.56 ± 0.03 | 5.77 ± 0.07 | 7.51 ± 0.16 | 3.68 ± 0.04 | F=385.543 | <0.001 |
| CVD, n (%) | | | | | | $\chi^2=147.811$ | <0.001 |
| No | 26300 (93.25) | 17629 (95.28) | 4958 (89.35) | 1633 (80.95) | 2080 (94.70) |  |  |
| Yes | 2415 (6.75) | 1067 (4.72) | 722 (10.65) | 484 (19.05) | 142 (5.30) |  |  |
| Status, n (%) | | | | | | $\chi^2=7.627$ | <0.001 |
| Survival | 1709 (75.60) | 811 (80.51) | 500 (74.29) | 295 (64.14) | 103 (76.05) |  |  |
| All-cause mortality | 706 (24.40) | 256 (19.49) | 222 (25.71) | 189 (35.86) | 39 (23.95) |  |  |
| Follow-up time, Mean ± SE | 69.98 ± 1.67 | 75.07 ± 2.51 | 68.57 ± 2.06 | 55.93 ± 2.49 | 76.62 ± 4.42 | F=9.842 | <0.001 |

F: ANOVA, $\chi^2$ : chi-square test.
SE: standard error, PIR: poverty income ratio, BMI: body mass index, MET: metabolic equivalent, DM: diabetes mellitus, Med: Mediterranean Diet, eGFR: estimated glomerular filtration rate, Na: sodium, K: potassium, BUN: blood urea nitrogen, CVD: cardiovascular disease.

### Association of serum osmolality with CVD

To investigate the relationship of serum osmolality with CVD in adults, we first screened the covariates associated with CVD (Table S1). Variables significantly linked to CVD (*P*<0.05) were included in the adjustment of multivariate models. Associations of hydration statuses with CVD in total participants are shown in the Table 2. After adjusting for covariates, we found that persons with current dehydration had higher odds of CVD compared with those with normal hydration (OR=1.26, 95%CI: 1.02-1.57).

**Table 2.** Associations of different hydration statuses with CVD.

| Hydration statuses | Unadjusted model |  | Model 1 |  | Model 2 |  |
| --- | --- | --- | --- | --- | --- | --- |
|  | OR (95% CI) | *P* | OR (95% CI) | *P* | OR (95% CI) | *P* |
| Normal hydration | Ref |  | Ref |  | Ref |  |
| Impending dehydration | 2.40 (2.13-2.72) | <0.001 | 1.52 (1.34-1.73) | <0.001 | 1.22 (1.06-1.41) | 0.007 |
| Current dehydration | 4.75 (3.89-5.80) | <0.001 | 2.36 (1.93-2.87) | <0.001 | 1.26 (1.02-1.57) | 0.033 |
| Hyperhydration | 1.13 (0.90-1.42) | 0.298 | 1.26 (0.99-1.60) | 0.060 | 1.15 (0.90-1.46) | 0.265 |

CVD: cardiovascular disease, OR: odds ratio, CI: confidence interval, Ref: reference.
Model 1: adjusted for age, gender, race, PIR, marital status, and education level;
Model 2: adjusted for age, gender, race, PIR, marital status, education level, BMI, physical activity, smoking, stroke, cancer, hypertension, DM, dyslipidemia, cardiovascular agents, total energy intake, and eGFR.

Additionally, we explored these relationships in different subgroups (Table 3). Current dehydration (serum osmolality >300 mmol/L) was linked to higher odds of CVD in ≥60 years old (OR=1.28, 95%CI: 1.01-1.61), male (OR=1.33, 95%CI: 1.00-1.76), non-stroke (OR=1.36, 95%CI: 1.08-1.71), non-cancer (OR=1.27, 95%CI: 1.01-1.59), hypertension (OR=1.31, 95%CI: 1.06-1.62), and non-DM (OR=1.42, 95%CI: 1.04-1.93) subgroups, compared with normal hydration (serum osmolality of 285-294 mmol/L). Besides, participants with impending dehydration (serum osmolality of 295-300 mmol/L) were seemed to have higher odds of CVD compared with those who with normal hydration in ≥60 years old, male, non-stroke, non-cancer, and hypertension subgroups (all *P*<0.05).

**Table 3.**
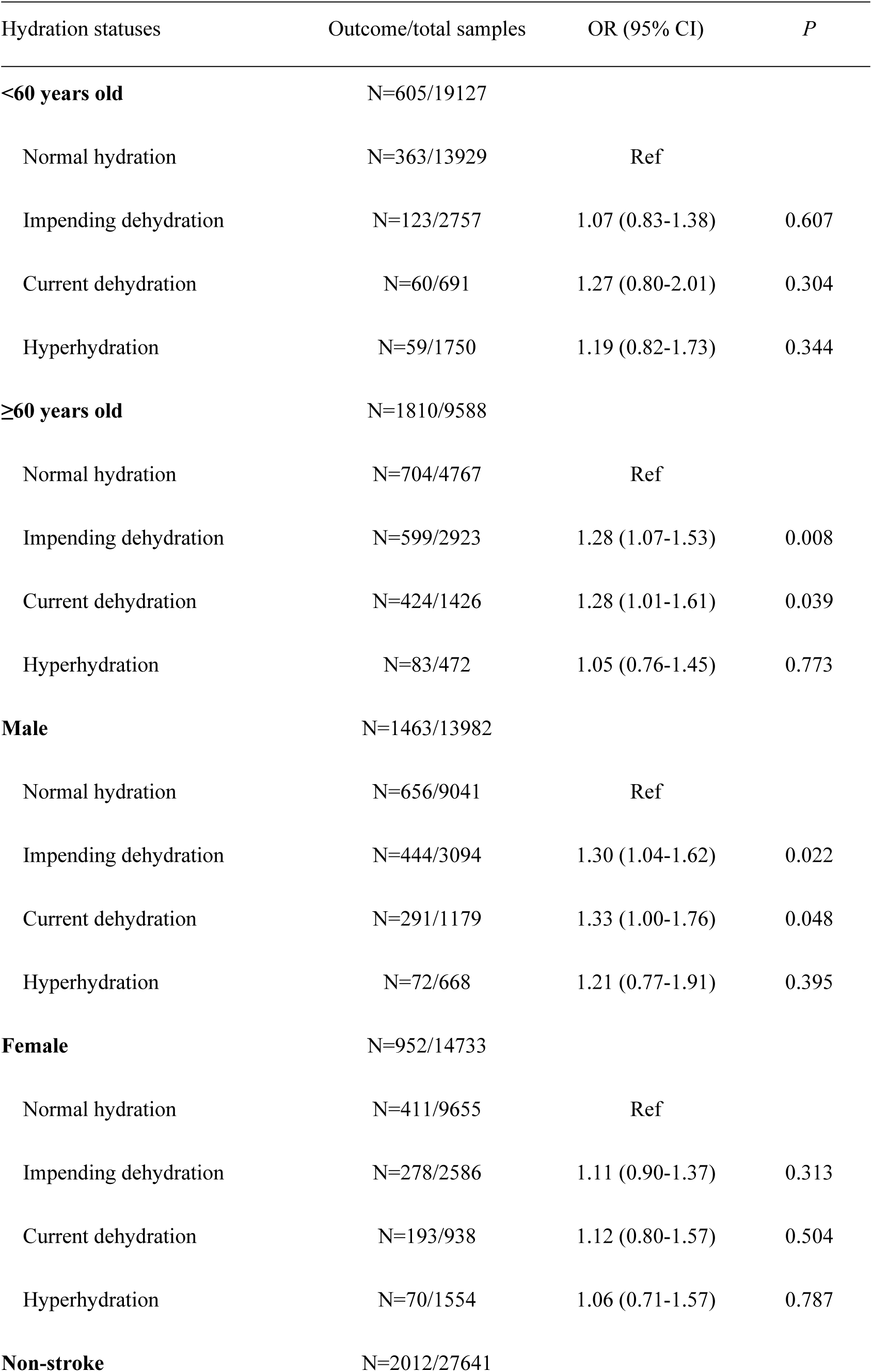
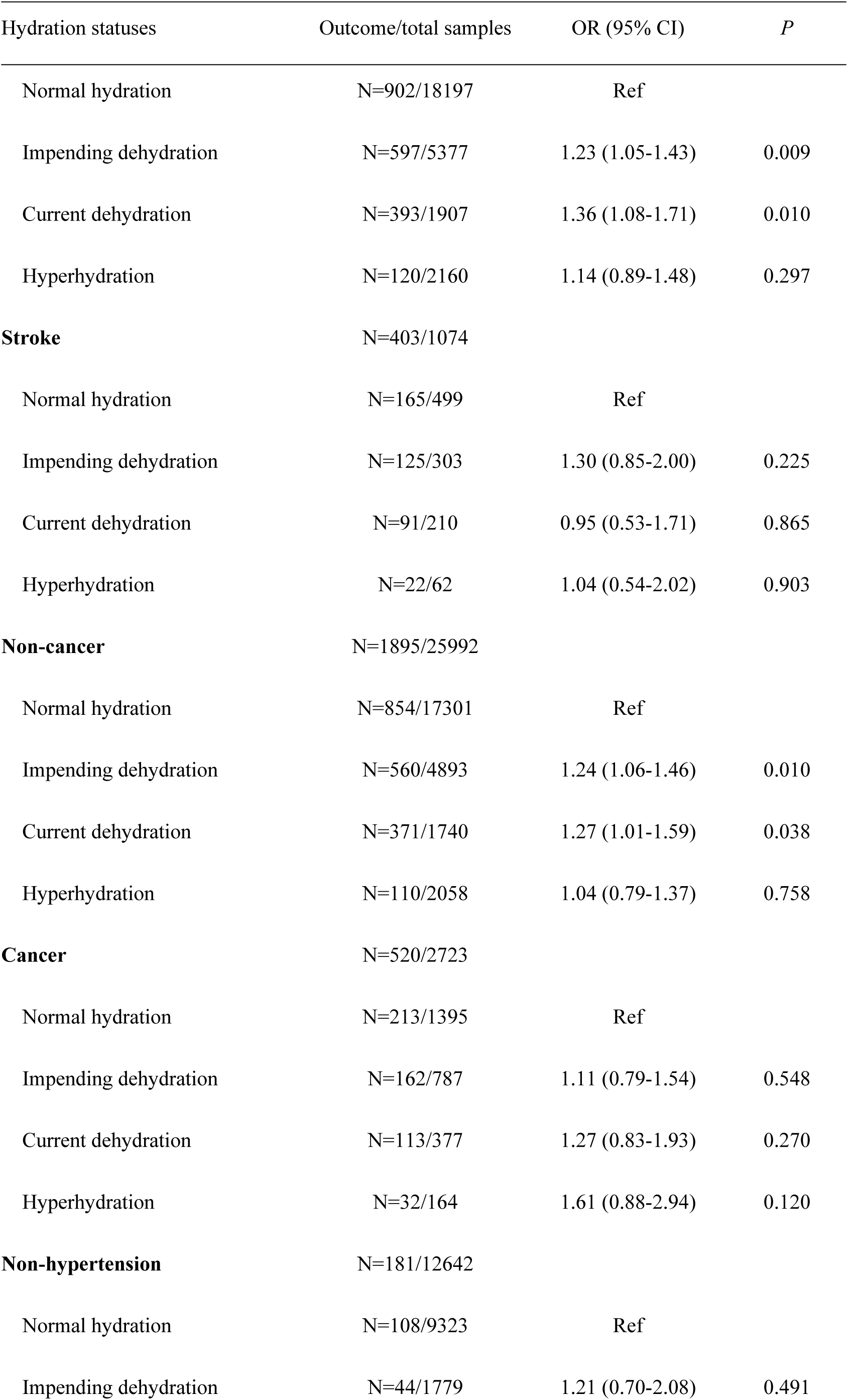
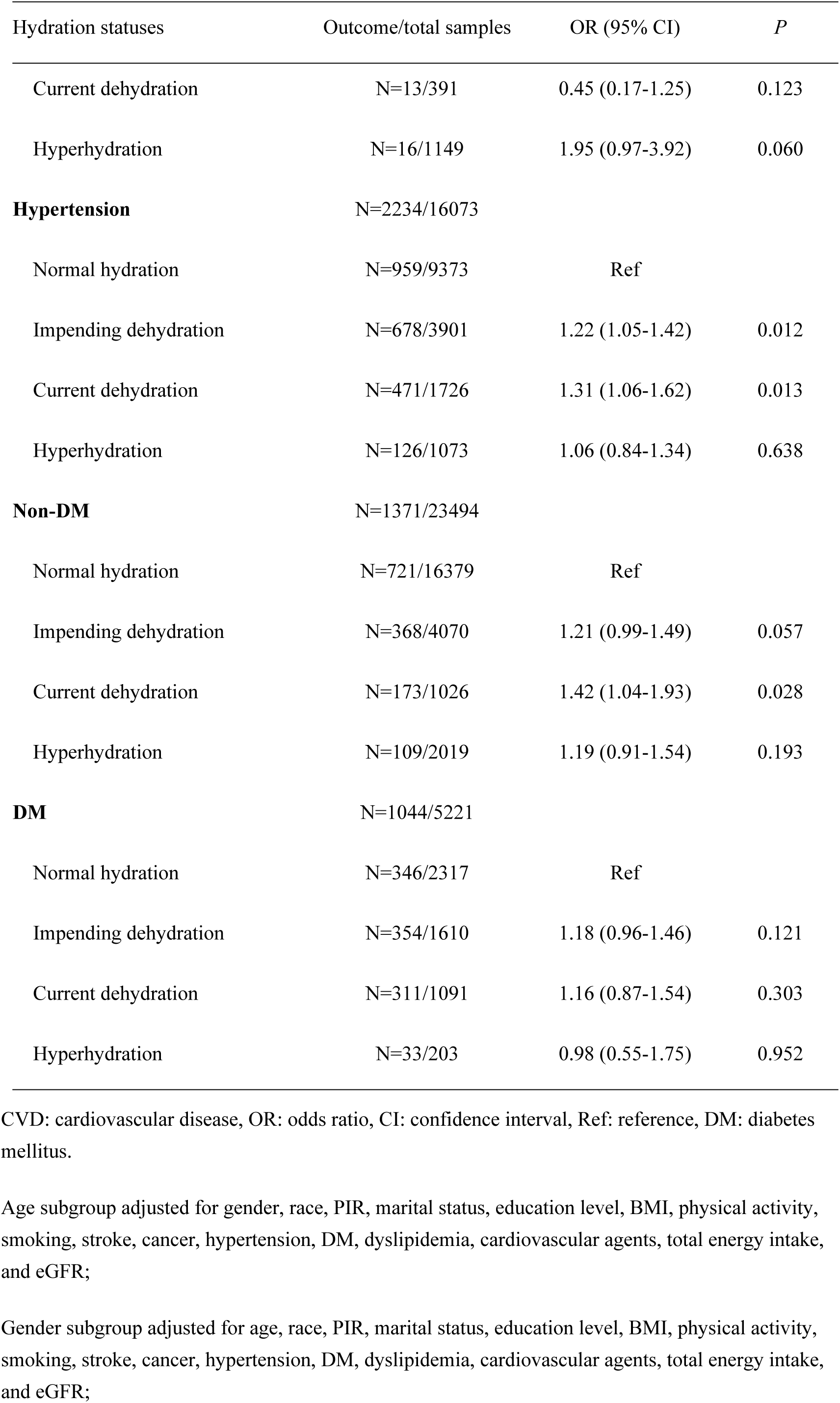
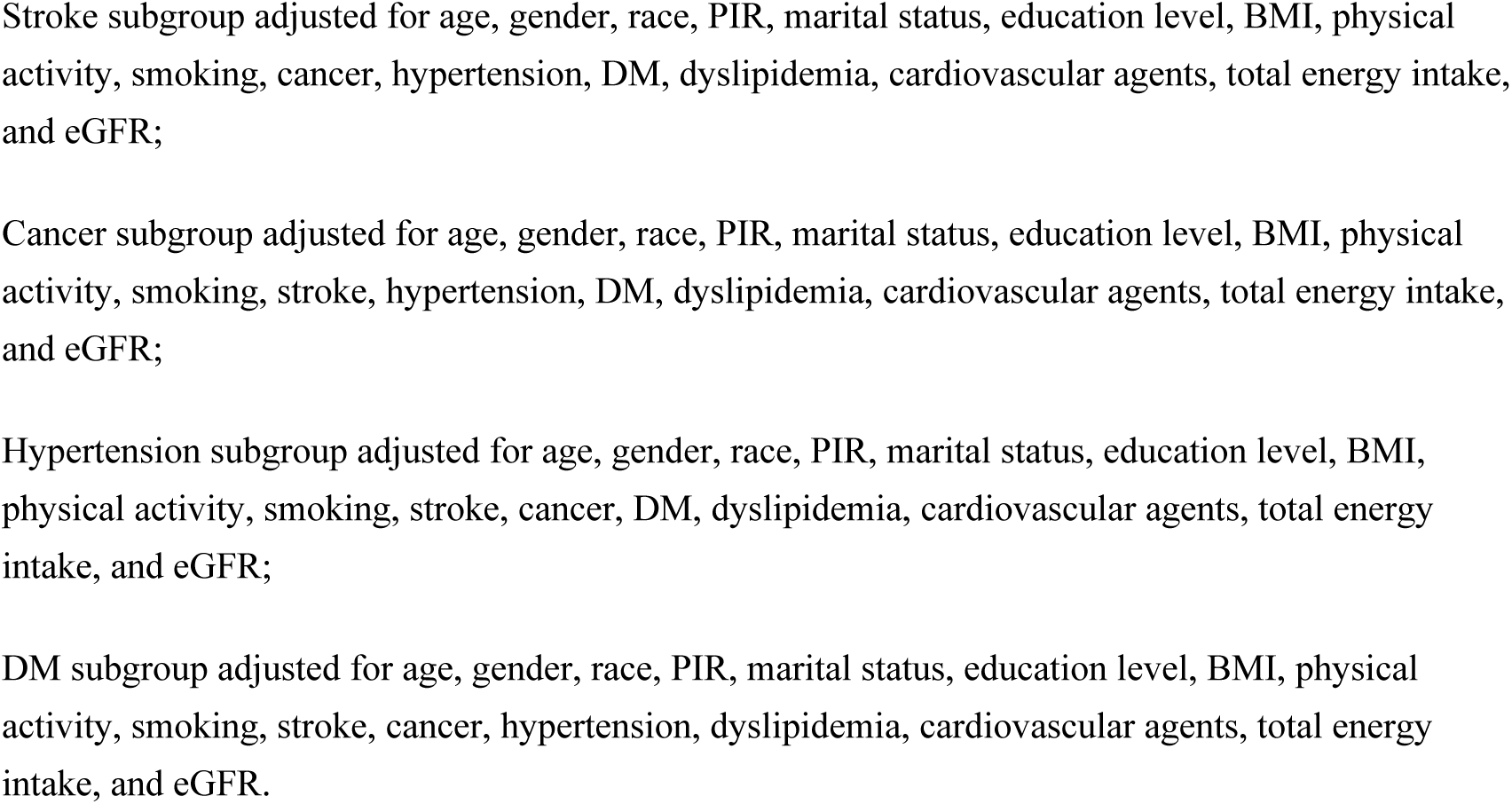
Associations of different hydration statuses with CVD in different subgroups.

### Relationship of serum osmolality with all-cause mortality in patients with CVD

We further assessed the association of different hydration statuses with all-cause mortality in persons who had CVD (Table 4). Similarly, we screened the covariates linked to all-cause mortality first (Table S2). Then, after adjusting for the selected covariates, patients with CVD who had current dehydration seemed to have higher risk of all-cause mortality compared with those had normal hydration (OR=1.41, 95%CI: 1.13-1.75). The KM curve also showed that among four hydration statuses, patients with CVD having current dehydration had the lowest survival probability, followed by those having impending dehydration (Figure 2).

**Figure 2.**
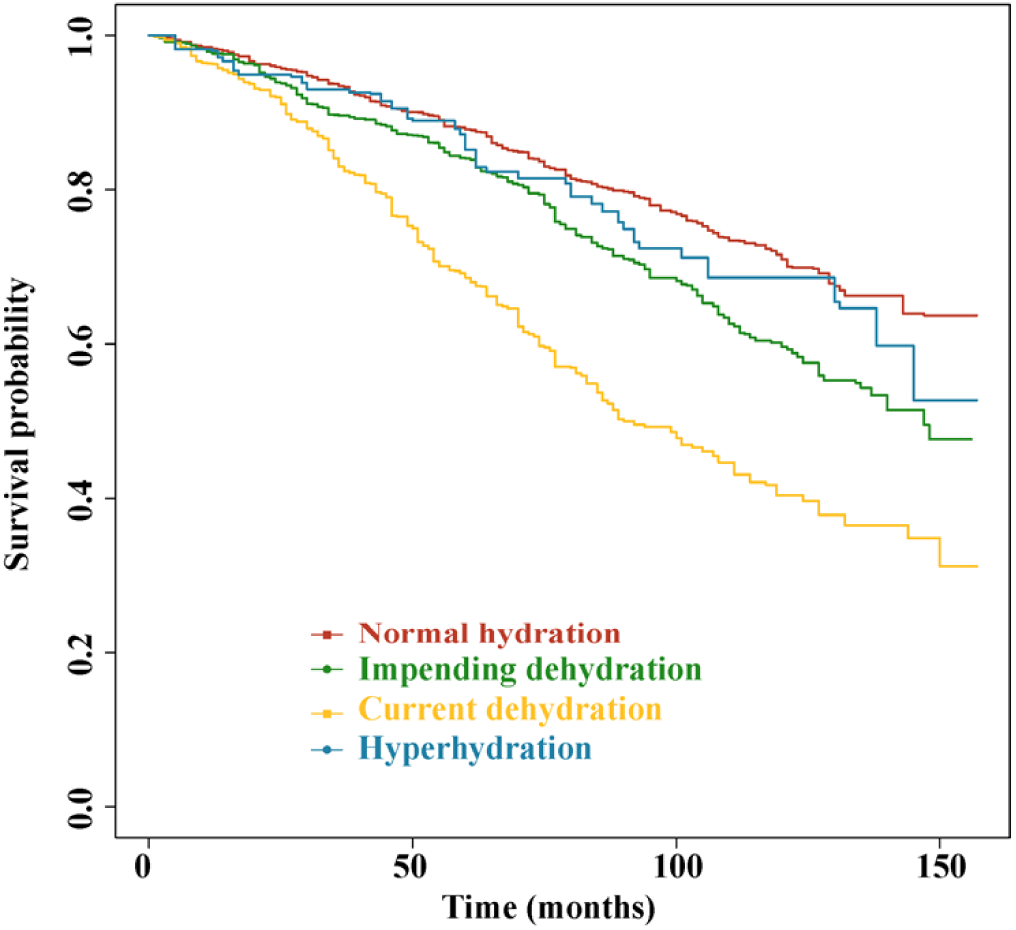
KM curves of different hydration statuses and survival probability in general populations.

**Table 4.** Associations of different hydration statuses with all-cause mortality in patients with CVD.

| Hydration statuses | Unadjusted model |  | Model 1 |  | Model 2 |  |
| --- | --- | --- | --- | --- | --- | --- |
|  | HR (95% CI) | *P* | HR (95% CI) | *P* | HR (95% CI) | *P* |
| Normal hydration | Ref |  | Ref |  | Ref |  |
| Impending dehydration | 1.48 (1.16-1.89) | 0.002 | 1.22 (0.95-1.56) | 0.120 | 1.03 (0.78-1.36) | 0.844 |
| Current dehydration | 2.69 (2.08-3.48) | <0.001 | 2.04 (1.57-2.65) | <0.001 | 1.41 (1.13-1.75) | 0.002 |
| Hyperhydration | 1.20 (0.72-2.01) | 0.479 | 1.18 (0.73-1.90) | 0.500 | 1.22 (0.72-2.07) | 0.453 |

CVD: cardiovascular disease, HR: hazard ratio, CI: confidence interval, Ref: reference.
Model 1: adjusted for age, race, marital status, and education level;
Model 2: adjusted for age, race, marital status, education level, physical activity, smoking, stroke, cancer, hypertension, DM, cardiovascular agents, aMed score, and eGFR.

In addition, these relationships were investigated in different subgroups (Table 5). Subgroup analyses results indicated that current dehydration was significantly linked to higher risk of all-cause mortality compared with normal hydration in ≥60 years old (OR=1.49, 95%CI: 1.19-1.87), female (OR=1.67, 95%CI: 1.14-2.44), non-stroke (OR=1.29, 95%CI: 1.01-1.64), stroke (OR=1.73, 95%CI: 1.01-2.97), non-cancer (OR=1.55, 95%CI: 1.17-2.06), hypertension (OR=1.43, 95%CI: 1.15-1.78), and DM (OR=1.50, 95%CI: 1.05-2.15) subgroups. Moreover, we observed that impending dehydration was linked to lower risk of all-cause mortality in CVD patients aged <60 years old (OR=0.29, 95%CI: 0.12-0.73), and hyperhydration was associated with higher risk of all-cause mortality in those who without cancers (OR=1.72, 95%CI: 1.04-2.85).

**Table 5.**
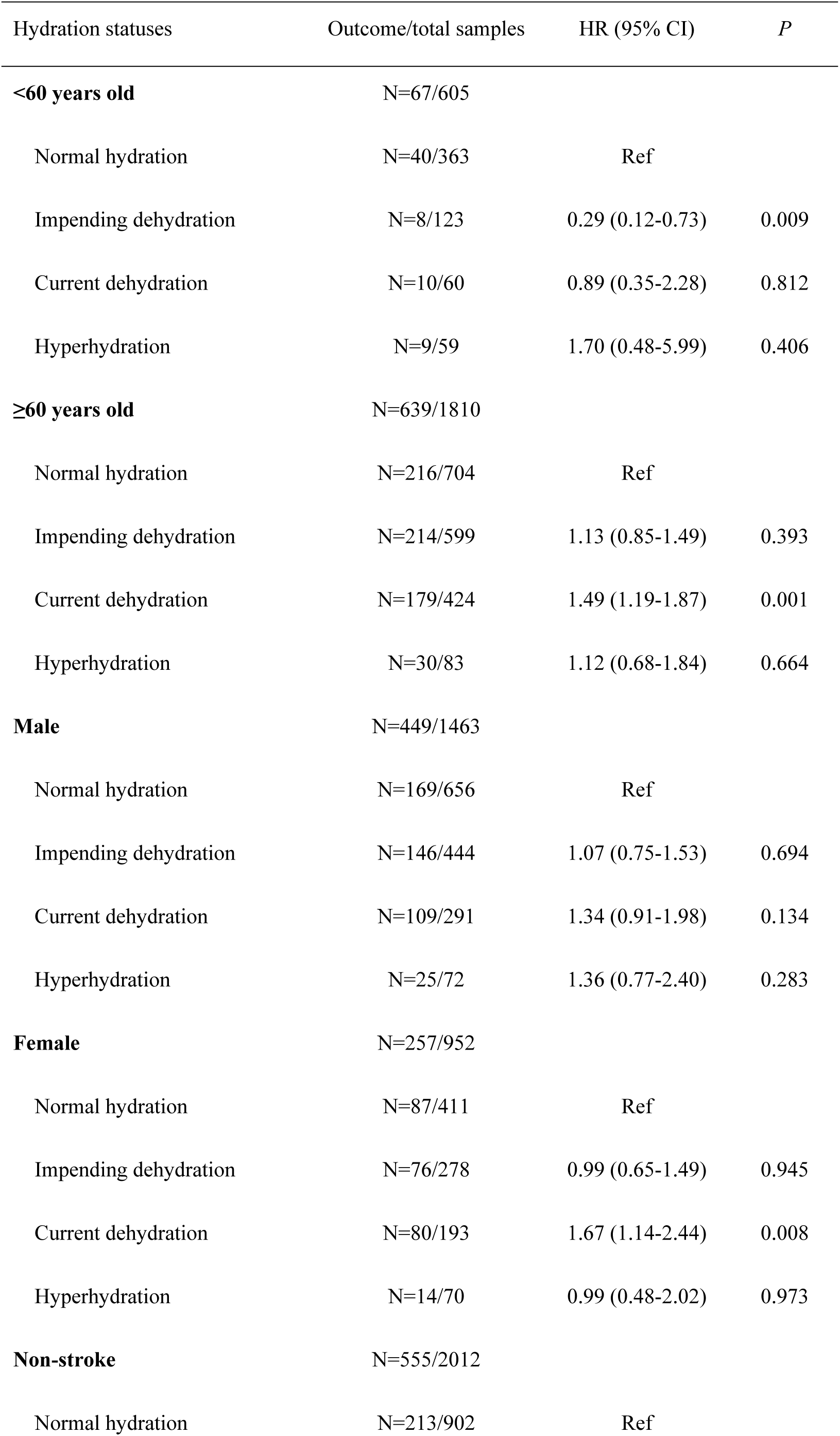
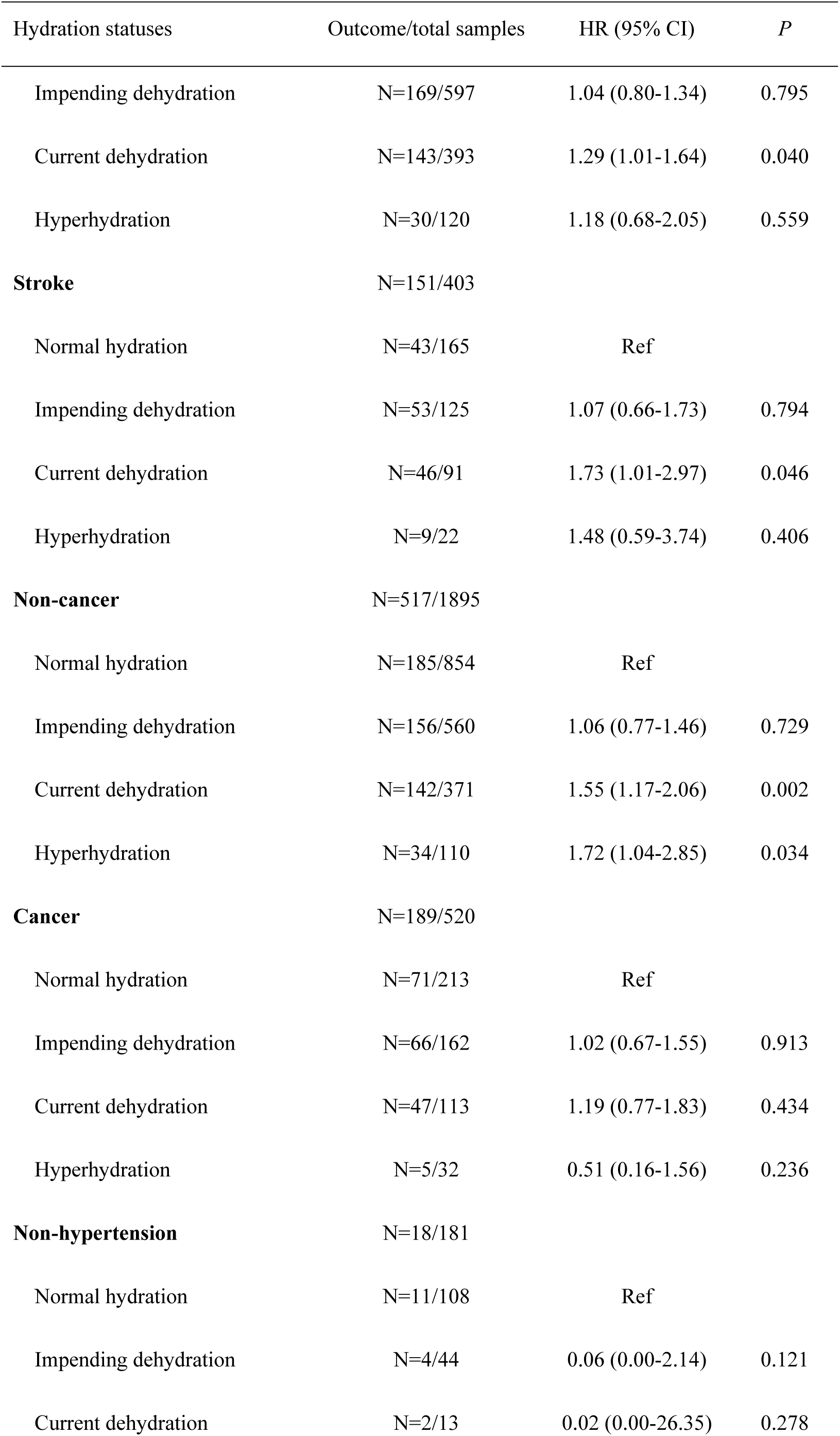
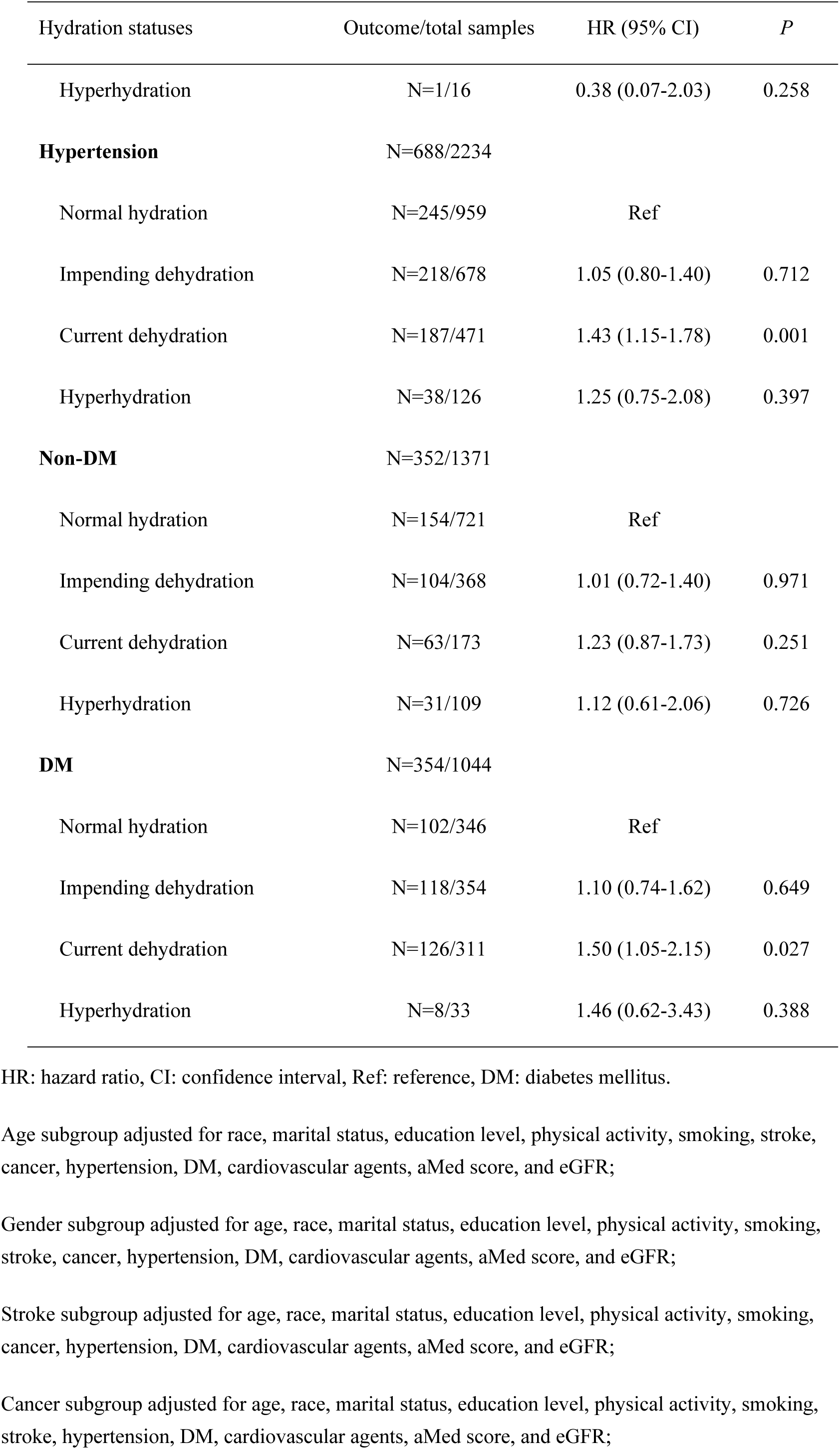
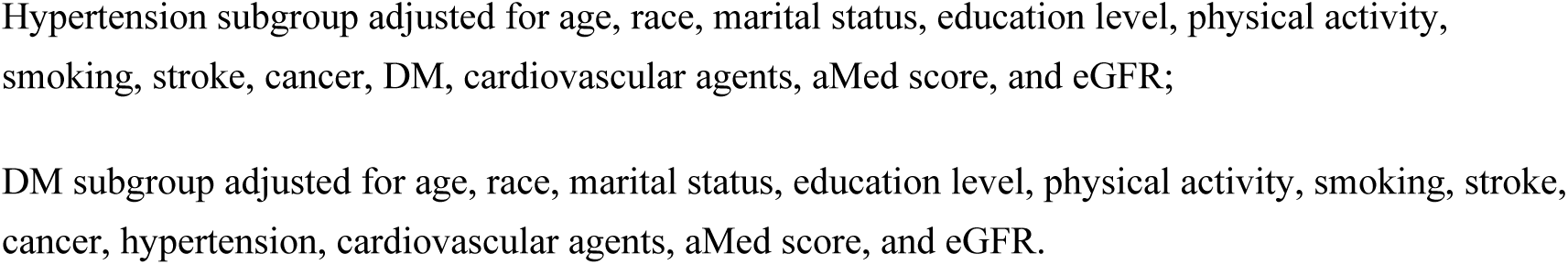
Associations of different hydration statuses with all-cause mortality in subgroups.

## Discussion

In the current research, we investigated the association of serum osmolality and CVD in adults, as well as the relationship of serum osmolality and all-cause mortality risk in patients with CVD. The study results showed that after adjusting for covariates, current dehydration was associated with higher odds of CVD in total population, and with higher risk of all-cause mortality in patients with CVD, compared with normal hydration. Also, the relationship between current dehydration and CVD were found in subgroups of ≥60 years old, male, non-stroke, non-cancer, hypertension, and non-DM.

No study has explored the role of serum osmolality in CVD among general population until now. A recent prospective cohort study produced by Wang et al. [12] investigated the association of serum osmolality with all-cause and cardiovascular mortality in adults in the United States, and found that serum osmolality was nonlinearly linked to all-cause and cardiovascular mortality, with an inflection point of 278 mmol/kg. Similar to Wang’s, this retrospective cohort study focused on the association of serum osmolality with CVD in general adults in the United States. We divided the serum osmolality into four levels and found that compared with normal hydration (serum osmolality of 285-294 mmol/L), impending dehydration (serum osmolality of 295-300 mmol/L) as well as current dehydration (serum osmolality >300 mmol/L) were both linked to higher odds of CVD, which seemed that this relationship was also nonlinearly. Our findings relatively filled in the literature blank in role of serum osmolality in CVD. In addition, Hu et al. [22] assessed relationship of serum osmolality with the risk of all-cause and cardiovascular mortality in patients with DM, where the restricted cubic splines results indicated that serum osmolality levels had a U-shaped association with the risk of all-cause mortality, and L-shaped relationship with the risk of cardiovascular mortality. In Hu’s report, the reference serum osmolality category was 281-284 mmol/kg, and the lowest and highest osmolality category were respectively <201 mmol/kg and >312 mmol/kg. It may be because the differences in the methods of hematology indicators used in the NHANES, so the specific cut-off values are somewhat different, but on the whole, our conclusions are relatively reliable. Nevertheless, the causal association of different hydration statuses with the risk of CVD needs further verification.

The underlying mechanisms that influence the relationship between osmolality and CVD are puzzling. Increased serum osmolality results in symptoms, such as osmotic diuresis, which can exacerbate dehydration and electrolyte imbalances. A prospective single center cohort study considered that serum osmolality acted as a predictor of adequate diuresis in acute decompensated heart failure [23]. In addition, a low osmolality shock increases the cell swelling, and enhance the cytocidal effects at the same time, which may lead to the cell shrinkage and rupture [24]. Aoki et al. [25] concluded that decreased serum osmolality promotes ductus arteriosus constriction, and a transient decrease in serum osmolality may promote ductus arteriosus closure during the first few days of life. However, we have not observed significant association of low serum osmolality with CVD in adults, and the possible mechanisms are also unclear. In fact, underhydration is a common condition in adults among the United States, which was cross-sectionally associated with the increased prevalence in obesity, metabolic syndrome, insulin resistance, DM, high waist circumference, hypertension, and low high-density lipoproteins [26]. Underhydration is associated with the chronic activation of the renin angiotensin system, endothelin, vasopressin, and the aldose reductase-fructokinase pathway that are involved in the developments of CVDs [27, 28]. However, the specific mechanisms that dehydration influencing CVD, and further mortality need further exploration.

We also performed these associations of different hydration statuses with CVD in subgroups of age, gender, stroke, cancer, hypertension, and DM. Results of subgroup analyses indicated that adults who aged ≥60 years old should pay great attention to hydration related serum indexes in clinical practice, especially the impending dehydration and current dehydration status. A recent study conducted over 95% of United States adults (aged 51-70 years old) were underhydrated, and there was a prevalence in elevated serum osmolality of 60% in older adults (≥295 mmol/L) [29, 30]. Among our study population, only 9,588 (26.32%) participants aged ≥60 years old. Similar to aging, the physiological responses to dehydration is affected by sex difference [31]. Leurs et al. [32] suggested higher total fluid consumption was not associated with either ischemic heart disease (IHD) mortality or stroke mortality in men or women. It was similar to our results that no significantly association between hyperhydration and CVD in neither male adults or female adults. However, the relationship of specific beverages consumption, such as coffee and tea with IHD mortality was opposite between females and males in Leurs’ study. As mentioned above, hydration statues are linked to various chronic diseases, and the subgroup analyses showed that individuals without stroke, cancer, or DM were recommended to dynamic monitoring serum indexes related to hydration status to reduce the potential risk of CVD. Differently, associations of impending dehydration and current dehydration with high odds of CVD were only found in patients with hypertension instead of those without hypertension. An imbalance of body Na and water can cause various pathological states, such as dehydration, hypertension, CVD, and metabolic disorders [33]. Among the four hydration statuses groups, persons with current dehydration had the highest serum levels of Na, K, glucose, and BUN, followed by those with impending dehydration. High Na status reduces heart rate and cardiovascular energy expenditure via suppression of renal sympathetic nerve activity to compensate for the energy-intensive water conservation, and that a high heart rate and a catabolic state such as muscle wasting and cachexia are major risk factors of cardiovascular events [34, 35]. In conclusion, this research provided some references for the prevention of CVD in the general population, and the risk of mortality in subsequent patients with CVD in clinical practice that serum osmolality is a potential convenient indicator.

One of the strengths of our research was the large and representative study sample extracted from the NHANES database. Also, the calculated serum osmolality was easy to access, and through investigating relationship of different hydration statuses with CVD and all-cause mortality in general population as well as different subgroups, we could provide the basis for risk stratification of specific population. However, some limitations limiting the interpretation on results. When exploring association of serum osmolality with CVD, the study design was cross-sectional study, which can not conclude a causal relationship. In addition, due to the limitation of the NHANES database, information on patients with CVD during the follow-up time was not available so that there may be unadjusted confounding factors.

### Conclusion

Calculated serum osmolality may be a potential indicator of CVD risk in general population because current dehydration was linked to high odds of CVD. Besides, among patients with CVD, dehydration status should also not be ignored, and clinicians need to adjust the dehydration in time to reduce mortality risk of patients.

## Additional Files

File name: Additional file

File format: .docx (Microsoft Word document)

Title: Supplementary Tables S1-S2

Description: This file contains supplementary tables (Table S1 and Table S2) that support the findings of this article.

## Availability of data and materials

The datasets generated and/or analysed during the current study are available in the NHANES repository,[https://www.cdc.gov/nchs/nhanes/index.htm].

## Data Availability

[https://www.cdc.gov/nchs/nhanes/index.htm].

## Acknowledgements

The authors thank the National Center for Health Statistics (NCHS) for providing the NHANES data and the participants who contributed to the survey.

